# Influences of race and clinical variables on psychiatric genetic research participation: Results from a schizophrenia sample

**DOI:** 10.1101/2022.11.03.22281884

**Authors:** Rose Mary Xavier, Yuktha Shanavas, Brian M. Britt, Wales T. George

## Abstract

**Objective:** Advances in genetics has led to a better understanding of both genetic and environmental contributions to psychiatric mental health disorders. But psychiatric genetics research is predominantly Eurocentric, and individuals of non-European ancestry continue to be significantly underrepresented in research studies. The objective of this study was to examine factors associated with genetic study participation in a schizophrenia sample.

**Methods:** The study sample was extracted from the Clinical Antipsychotics Trial of Intervention Effectiveness (CATIE) schizophrenia study which enrolled patients with schizophrenia between the ages of 18-65 and incorporated an optional genetic sub-study. Using regression models, we examined sociodemographic and clinical factors that were independently associated with participation in the genetic sub-study.

**Results:** The genetic sub-study had a lower proportion of Black (30% in genetic vs 40% in CATIE overall) and Other race (4% vs 6%) participants. Severe psychopathology symptoms (odds ratio [OR]=0.78, p=0.004) and better reasoning scores (OR = 1.16, *p* = 0.036) influenced the odds of genetic study participation. Compared to Black participants, White participants were significantly more likely to participate in the genetic sub-study (OR=1.43, p=0.009)

**Conclusion:** Race and clinical variables significantly impact genetic study participation of individuals with schizophrenia. Future studies should examine the interactive effects of race and clinical variables for a nuanced understanding of how individuals diagnosed with severe psychiatric illnesses choose to participate in genetics studies.

## Introduction

Genetic studies have overwhelmingly focused on individuals of European ancestry (Atkinson et al., 2022; Peterson et al., 2019; Popejoy & Fullerton, 2016). Despite concerted efforts to increase representation, individuals of African, Latin American, Hispanic, Indigenous and East Asian ancestry continue to be severely underrepresented in genetic studies (Popejoy & Fullerton, 2016). For genetic studies of psychiatric and mental health disorders, this underrepresentation is quite pronounced (Martin, Daly, et al., 2019; Martin, Kanai, et al., 2019). Scientific insights gained from one population does not carry over to another as differences in allele frequencies and genetic background create significant confounds (Freedman et al., 2004). Underrepresentation of individuals of non-European ancestry in psychiatric genetic studies has the potential to worsen existing mental health disparities (Martin, Kanai, et al., 2019). Understanding factors that determine research participation is important to facilitate improved participation in psychiatric genetic studies and is critically important for underrepresented patient participants.

Patients’ research priorities often don’t align well with those of researchers and funding agencies (Crowe et al., 2015). What patients’ perceive as important can influence their research participation levels. Studies that have specifically examined factors affecting psychiatric and mental health research participation have identified fear of invasive procedures (Schafer et al., 2011), the type of research (e.g., genetic studies, neuroimaging etc.), a lack of comprehension regarding research topic and study procedures, fear of treatment side effects, study safety and social stigma surrounding mental health illnesses as major factors that influence participation (Murphy & Thompson, 2009; Roberts & Kim, 2017). The specific psychiatric diagnosis of the participant is also associated with their research participation. For example, compared to patients with depression, patients with schizophrenia report a lower approval of psychiatric research and a decreased readiness to participate in research using questionnaires and those requiring blood draws (Schafer et al., 2011). For both diagnostic groups, there is a lower level of willingness to participate in studies that are medication trials or that use neuroimaging techniques (Schafer et al., 2011). Patients with schizophrenia generally do not positively regard research if the methods were perceived as dangerous and involuntary (Weiss Roberts et al., 2000).

Studies that have examined factors that impact research participation in psychosis and schizophrenia have primarily focused on the motivations and incentives that increase research participation. These studies show that the main factors incentivizing a schizophrenia patient’s participation included altruism (i.e., helping others and science) (Weiss Roberts et al., 2000; Zullino et al., 2003) and monetary rewards (Schafer et al, 2011); how participants were informed and referred to research studies also influence research participation among patients with psychosis. For example, clear communication and a positive patient-clinician relationship were both key factors that increased research participation (Bucci et al., 2015). When the patient-clinician relationship is generally positive, patients were more likely to be referred to psychosis research studies (Bucci et al., 2015).

Sociodemographic characteristics can influence who are exposed to opportunities for research participation as well as who are likely to participate in research studies. For example, gender and minority status influence how and where an individual access psychiatric care which in turn can influence recruitment into research studies (Brown et al., 2014). Black and other persons of color experience multiple obstacles to active engagement in research including time constraints, lack of health insurance, lack of media access, and transportation issues to and from research sites (Hussain et al, 2004; Murphy & Thompson, 2009). Black participants’ also have a higher level of mistrust regarding participation in genetic research compared to White participants regardless of their socioeconomic status and education levels (Furr, 2002; Laskey et al., 2003; McQuillan et al., 2003; Murphy & Thompson, 2009). Black participants report concerns about the safety behind procedures as well as confidentiality about mental illness in families (Brown et al., 2014; Murphy & Thompson, 2009). There is a dearth of knowledge regarding the social, demographic, and clinical factors associated with genetic research participation in the severely mentally ill such as those with schizophrenia and related psychosis.

The purpose of this study was to examine differences in characteristics of schizophrenia patients who chose to participate in a genetic study compared to those who chose not to. Specifically, we examined sociodemographic and clinical factors that were independently associated with participation in a genetic study. We also investigated differences in longitudinal trajectories of symptoms and clinical outcomes between the participants who donated the DNA sample for the genetic study compared to those who did not.

## Methods

We obtained our study sample from the Clinical Antipsychotics Trial of Intervention Effectiveness (CATIE) schizophrenia study conducted between 2001 and 2004. Briefly, CATIE was a multiphase randomized controlled trial (N = 1493) that compared effectiveness of antipsychotics medications in patients with chronic schizophrenia between the ages of 18-65. CATIE also incorporated an optional genetic sub-study that aimed to identify genetic risk variants associated with schizophrenia; -51% (N = 738) of the CATIE sample participated in the genetic study. Details of the parent study is available at Stroup et.al (2003). We accessed CATIE data from the NIMH data archive (https://nda.nih.gov/) after obtaining relevant data access and IRB approvals.

### Measures

*Genetic study participation* was measured based on if the participant donated a sample for the genetic sub-study-a dichotomous variable coded as ‘1’ for ‘Yes’ and ‘0’ for ‘No’. *Psychopathology symptoms* were rated using the positive and negative syndrome scale (PANSS) which has a total of 30 items with each item scored between 1 (absent) to 7 (extreme); the higher the scores the more severe the symptoms with total scores of <58 indicating mild illness, 58-75 as moderately ill, 76-95 markedly ill and 96-116 as severely ill. PANSS has 3 subscales - 7 items assessing positive symptoms, 7 negative symptoms and 16 items focused on general psychopathology symptoms. *Insight* was measured by the insight and treatment attitudes questionnaire (ITAQ), a 11-item scale measuring the participant’s attitude towards treatment and their awareness of illness. ITAQ total scores range from 0 (indicating no insight) to 22 (indicating full awareness of illness), with higher scores indicating better insight. *Symptom severity* was assessed from both the patient and the clinician perspectives using the *Clinical global impression of symptom severity* (CGIS)-the patient version measures the participant’s impression of their symptom severity. The clinician version is rated by clinicians to measure the global symptoms severity. Both items are scored from 1 to 7, with 1 being “Normal, Not ill” and 7 being “Very Severely Ill”. Severity of *depressive symptoms* in schizophrenia patients was measured by the *Calgary depression rating scale* (CDRS) which measures 9 mood symptoms which are scored from 1-4: less overall score indicates fewer depression symptoms. *Neurocognition* measure was a composite score derived from working memory, verbal memory, processing speed, reasoning, and vigilance domain scores. Please refer to Keefe et al. (2006) for details on how the composite scores were calculated. *Health summary* was measured by the SF-12, a 12-item participant reported survey of mental and physical health. We used physical and mental health summary variables in our study; these variable scores range from 0 to 100. Higher scores indicate better general physical and mental health with scores <50 used for physical summary to determine a physical condition and a score of <42 indicating depression. Patient symptoms and functioning over the preceding 4 weeks are measured using the *Quality of Life Scale* (QLS) (Heinrichs et al. 1984). It consists of 21 items, each rated on a 7-point scale and primarily relying on clinician judgment.

The *MacArthur Competence Assessment Tool-Clinical Research* (MacCAT-CR) is used to measure the capacity of the potential research participant to make competent decisions about research participation. We used 4 subscales in our analysis. (1) Understanding-the sum of the 13 item subset of the MacCAT-CR which pertains to a participant’s ability to understand provided information about a research project and its protocols. Scores can range from 0-26 with <= 15 suggests poor understanding. (2) Appreciation-the sum of the 3 items, can range between 0-6 and pertains to a participant’s ability to evaluate the consequences of study participation on themselves. (3) Choice is determined from the participant’s response to the item that pertains to a patient’s ability to clearly communicate a decision about their willingness to participate or not to participate in a research study. Scores can range between 0-2. (4) Reasoning is the sum of the 4 items with scores that can range between 0-8 and pertains to the patient’s ability to reason about their participation in a study by comparing alternatives and their consequences on themselves. *Medication adherence* was measured by the clinicians’ judgement on a scale of 1 to 4, with 1 being adherent always/most of the time, 2 for usually, 3 for sometimes and 4 for never/almost never. This was reverse coded for analysis.

### Statistical Analysis

We first tested for assumptions of normality by examining data distributions of all variables included in our analysis. Variables in our analysis were all near normally distributed. Since there were missing data, we tested the missing mechanism using the R package *naniar* (Tierney & Cook, 2018). The data did not meet the assumption of missing completely at random (χ = 450, df = 292, *p* <0.001) and subsequently we performed multiple imputation to estimate missing data using the R package *MissForest* (Stekhoven, n.d.). We conducted analysis on both the datasets - first on the original data with missing values and a second on the imputed dataset. To examine if socio-demographic (age, gender, race) and clinical variables (psychopathology symptoms, insight, depression, neurocognition, quality of life, capacity for decision making and antipsychotic medication) that were independently associated with genetic study participation, we estimated a logistic regression model with blood sample donation status for genetic sub-study participation as the outcome variable using baseline/study entry data. To investigate if there were longitudinal changes in clinical variables between the overall and genetic sub-study groups, we estimated a linear mixed model, assuming participant level random effects, with relevant clinical variables (psychopathology symptoms, insight, depression, neurocognition, general health, quality of life, capacity for decision making, illness severity) as the outcome variable and blood sample donation as the dependent variable adjusting for the effects of age, gender, race, and medication adherence across the study period.

## Results

Our overall study sample were 74% male and included 60% White participants, 35% Black participants and 5% were from other races-0.5% American Indian/Alaska Native, 2.3% Asian, 0.3% Hawaiian or Pacific Islander, 1.8% of mixed race, 0.1% unknown or not reported. Since the sample size for groups other than White and Black participants were small, we collapsed them into an ‘Other’ group for all analysis. Descriptive summaries of other variables are provided in table 1. 52.4% of the overall sample participated in the genetic sub-study. Differences between groups that participated in the genetic study compared to those who chose not to are provided in Table 2. The genetic sub-study had a lower proportion of Black (30% in genetic sub-study vs 40% in CATIE overall) and Other (4% vs 6%) participants.

**Table 1:** Descriptive summary of variables at baseline

| Variables | Mean(SD) | Range | Interquartile range |
| --- | --- | --- | --- |
| Age in years | 40.5 (11.1) | 18–67 | 17 |
| PANSS | 75.9 (17.7) | 31–140 | 23 |
| Insight | 18.1 (5.1) | 0–22 | 7 |
| Depression | 4.5 (4.4) | 0–22 | 6 |
| Neurocognition (standardized) | 0 (1) | -3.3–2.9 | 1.3 |
| General physical health | 48.2 (10.1) | 13–68 | 14.5 |
| General mental health | 40.9 (11.7) | 8–67 | 19 |
| Quality of Life | 2.7 (3.1) | 0.2–5.9 | 1.5 |
| Competency |  |  |  |
| Understanding | 23.4 (3.1) | 0–26 | 4 |
| Appreciation | 4.5 (1.3) | 0–6 | 2 |
| Reasoning | 5.7 (2) | 0–8 | 3 |
| Choice | 2 (0.1) | 0–2 | 0 |

**Table 2:**
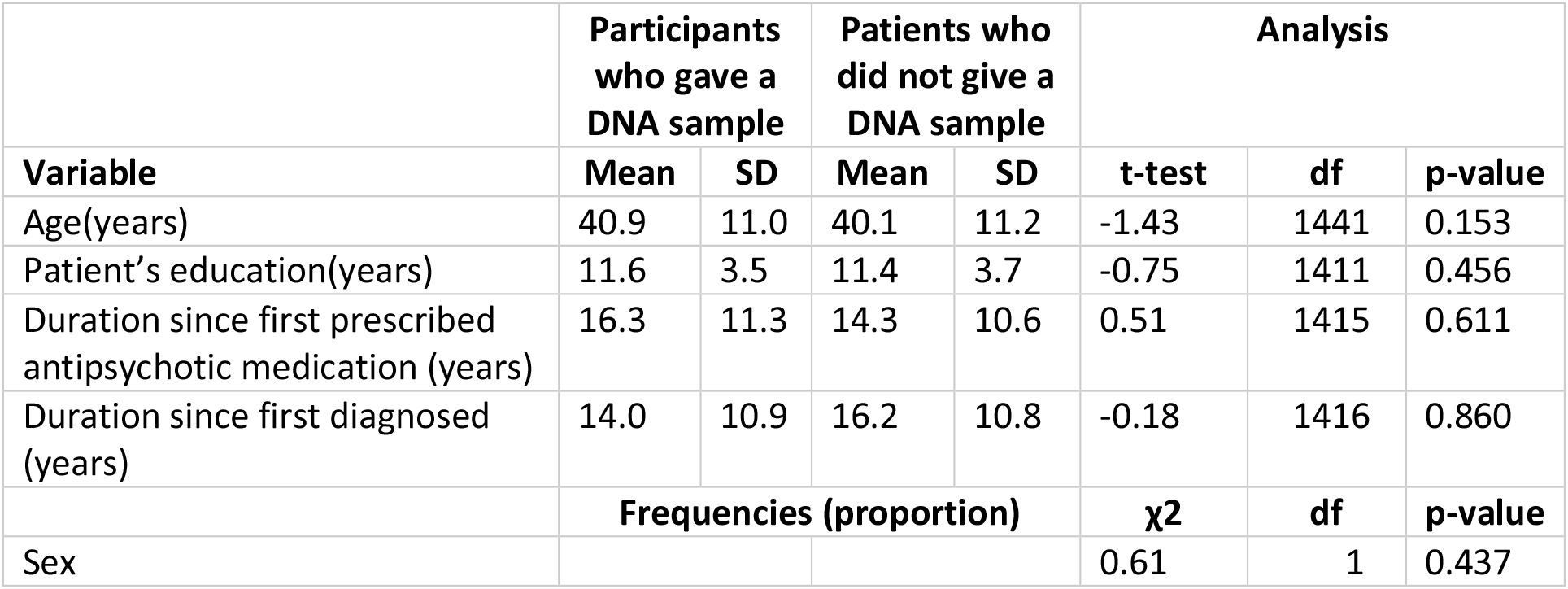
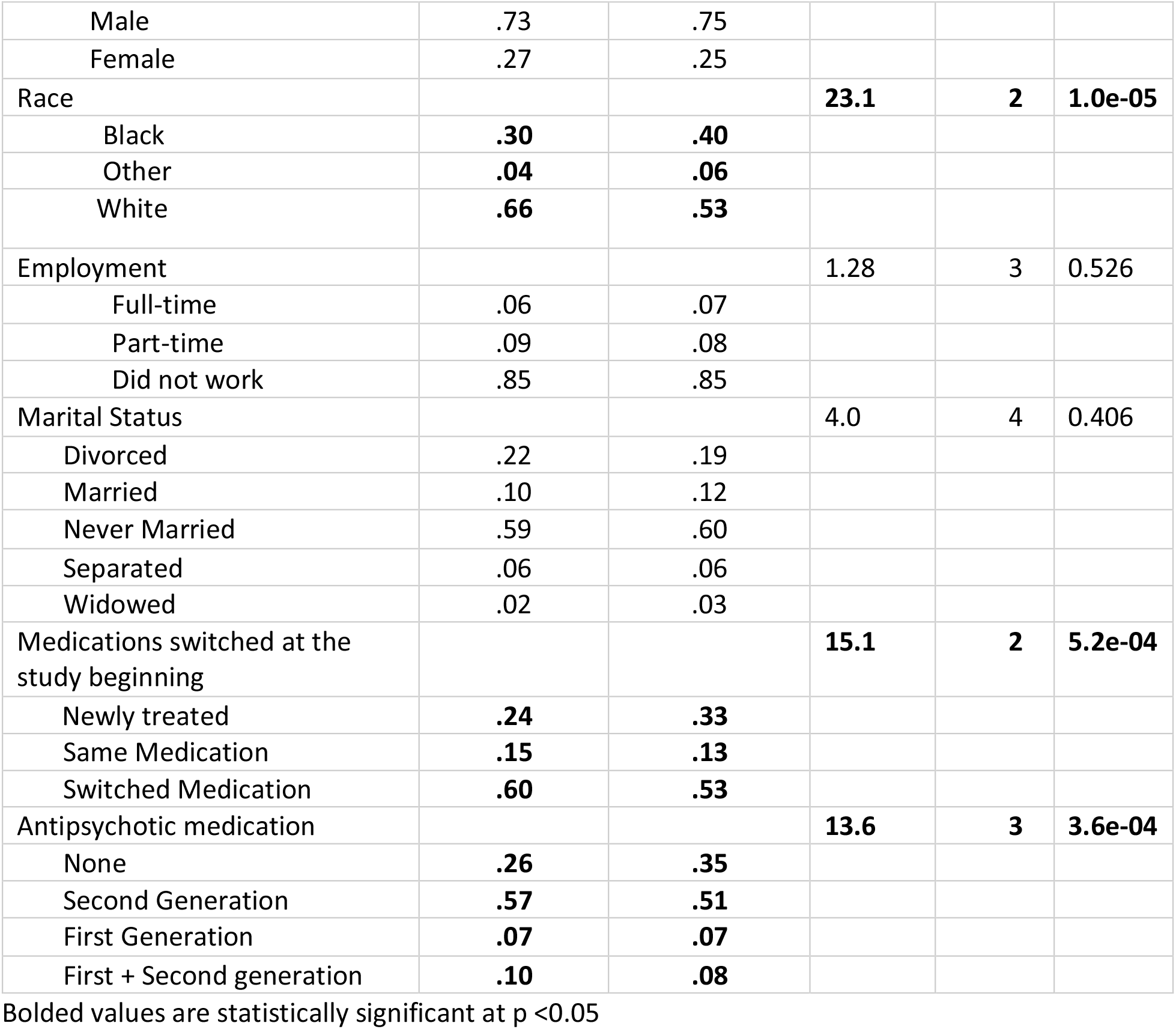
Socio-demographic characteristics by genetic sub-study participation

**Table 3:** Logistic regression results for baseline data (N = 1249)

|  | Variable | Complete data |  |  |
| --- | --- | --- | --- | --- |
|  |  | Standardized Coefficient (log odds) | 95% CIs | p-value |
| Sociodemographic variables | Age | 0.06 | -0.07, 0.19 | 0.350 |
|  | Gender (ref = female) | -0.04 | -0.31, 0.24 | 0.787 |
|  | Education in years | 0.01 | -0.03, 0.04 | 0.743 |
|  | Race (ref = Black) |  |  |  |
|  | **White** | **0.39** | **0.13, 0.64** | **0.003** |
|  | Other | -0.01 | -0.57, 0.55 | 0.967 |
| Clinical variables | Antipsychotic medication at baseline (ref = no med) |  |  |  |
|  | **2<sup>nd</sup> generation** | **0.34** | **0.07, 0.61** | **0.013** |
|  | 1<sup>st</sup> generation | 0.16 | -0.33, 0.65 | 0.522 |
|  | 1<sup>st</sup> + 2<sup>nd</sup> generation | 0.35 | -0.08, 0.78 | 0.110 |
|  | Insight | 0.05 | -0.07, 0.17 | 0.418 |
|  | Depression | 0.09 | -0.03, 0.22 | 0.147 |
|  | **PANSS total** | **-0.26** | **-0.40, -0.12** | **<0.001** |
|  | Neurocognition | -0.07 | -0.21, 0.07 | 0.308 |
|  | Quality of life | -0.10 | -0.25, 0.04 | 0.162 |
|  | MacArthur Competencies |  |  |  |
|  | Understanding | -0.12 | -0.25, 0.009 | 0.068 |
|  | Appreciation | 0.05 | -0.09, 0.19 | 0.510 |
|  | **Reasoning** | **0.15** | **0.01, 0.29** | **0.036** |
|  | Choice | 0.07 | -0.05, 0.19 | 0.246 |

Bolded values are statistically significant at p <0.05

**Table 4:** Longitudinal changes in clinical variables by genetic sub-study participation

| Outcome Variable | N | Coefficient<br>for sample<br>provided<br>(ref = No) | p-value |
| --- | --- | --- | --- |
| Insight | 1309 | **0.63** | **0.011** |
| Illness severity, patient rating | 1316 | -0.09 | 0.198 |
| Illness severity, clinician rating | 1316 | **-0.24** | **1.61e-07** |
| Depression | 1319 | 0.07 | 0.714 |
| PANSS | 1320 |  |  |
| Total |  | **-5.43** | **1.18e-09** |
| Positive |  | **-1.66** | **4.15e-09** |
| Negative |  | **-1.15** | **1.71e-04** |
| General |  | **-2.64** | **8.54e-09** |
| Neurocognition | 1141 | -0.05 | 0.372 |
| General physical Health | 1307 | -0.37 | 0.490 |
| General mental Health | 1307 | 0.20 | 0.707 |
| Quality of life | 1305 | 0.04 | 0.467 |
| MacArthur Competency<br>Subscales | 1230 |  |  |
| Understanding |  | -0.21 | 0.153 |
| Appreciation |  | 0.00 | 0.998 |
| Reasoning |  | 0.04 | 0.626 |
| Choice |  | 0.01 | 0.203 |

Gender, age, race, medication adherence and medication adherence x time (interaction) were included as covariates in all models; these estimates are not provided in the table.

For baseline data, the proportion of missing data for all variables of interest expect neurocognition were less than 2%; neurocognition was missing for -9% of the sample. We fitted a logistic regression model using maximum likelihood estimation to examine which sociodemographic and clinical variables at baseline were independently associated with genetic study participation. 95% confidence intervals and p-values were computed using the Wald approximation. Since the estimates for both the complete and imputed data were similar, we report and interpret our results from the complete data. Please see table 2 for full results of the model. Our results show that increased severity of psychopathology symptoms assessed by PANSS scores significantly decreased the odds of whether a participant gave a DNA sample (odds ratio [OR] = 0.77, *p* = <0.001). A participant’s increased level of reasoning competency increased the odds of them giving a sample (OR = 1.16, *p* = 0.036). Compared to Black participants, White participants were more likely to give a blood sample for genetic study participation (OR = 1.43, *p* = 0.003). Please see figure 1 for the predicted probabilities of genetic study participation for varying levels of psychopathology symptoms by race. Compared to participants who were not on any medications at baseline, those who on antipsychotic medications were more likely to provide a blood sample for genetic sub-study participation. But this association was only statistically significant for second generation antipsychotic medications (OR = 1.41, *p* = 0.013). Other variables included in the model were not statistically significant.

**Figure 1:**
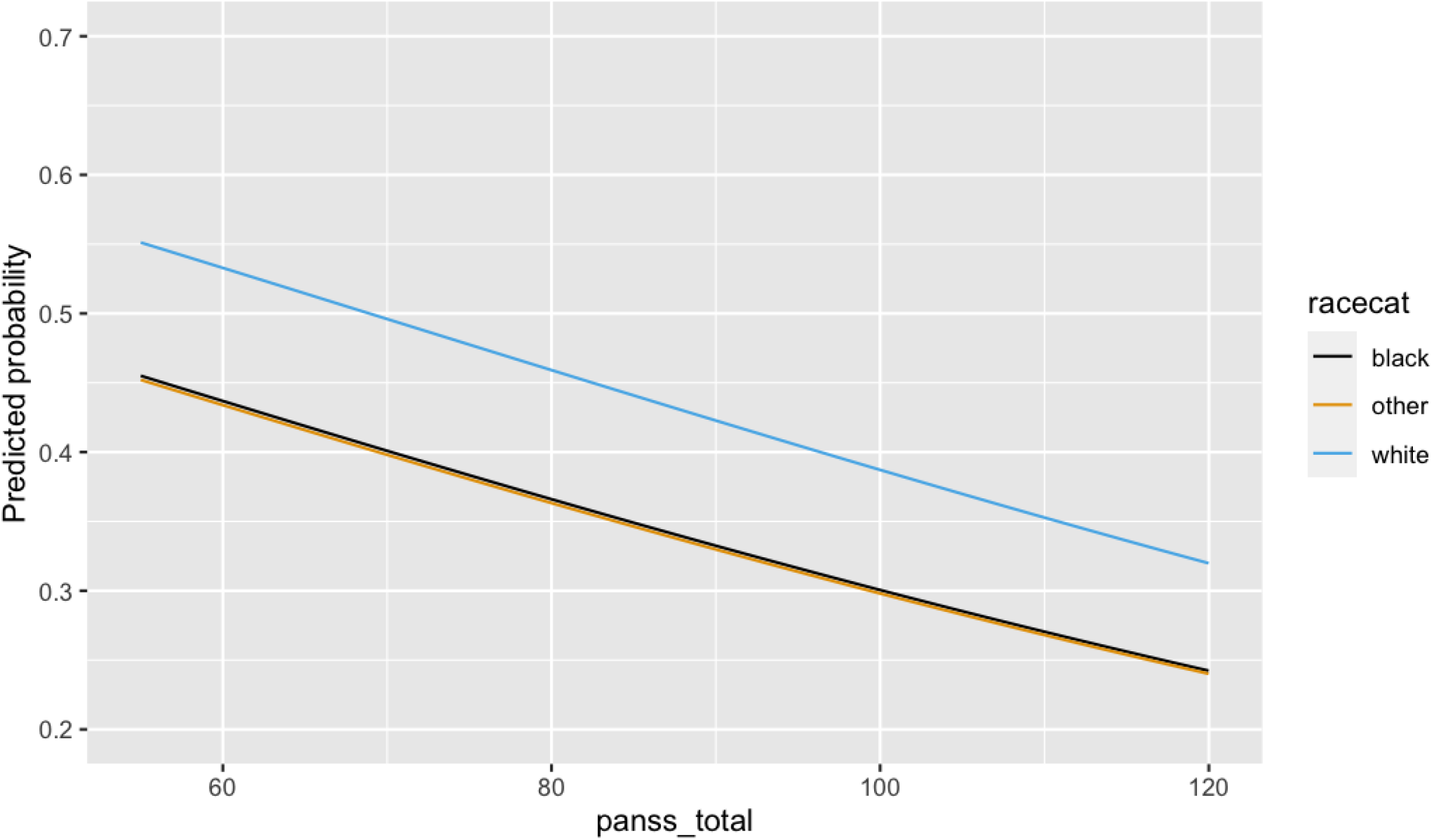
Predicted probabilities of genetic study participation from the logit model. Y-axis indicates probabilities and x-axis indicates psychopathology symptom scores measured by PANSS. At any level of psychopathology symptoms, Black and Other participants were less likely to participate in the genetic study compared to White participants.

Missing data proportions for longitudinal data are provided in the supplementary Table 1. We selected only those timepoints where data was consistently collected for a variable on most participants. We fitted linear mixed models using restricted maximum likelihood estimation on both the original dataset and imputed dataset to examine longitudinal changes in clinical variables grouped by genetic study participation (Table 4). Estimations from original data were very similar to imputed data estimations, so we report and interpret results only from the original data. Compared to others, participants in the genetic sub-study longitudinally had significantly better insight (*β* = 0.63, *p* <0.05), were rated by clinician as less severely ill (*β* = -0.24, *p* <0.001) and had less severe psychopathology symptoms (*β* = -5.43, *p* <0.001) across the study period.

## Discussion

Our study examined sociodemographic and clinical characteristics of genetic study participation using a relatively large sample of individuals with chronic schizophrenia. We report that among individuals with chronic schizophrenia, clinical factors (severity of psychopathology symptoms, reasoning abilities and illness severity in general) and sociodemographic factors such as self-reported race significantly influence genetic study participation. Our finding that individuals with severe symptoms and poorer reasoning scores (which are strongly correlated with cognitive dysfunction in schizophrenia; please refer to supplementary figure 1) were less likely to participate presents a challenge for investigations into the genetic contributors to schizophrenia. Individuals with the most severe illnesses can be highly informative samples not just to understand illness etiology, but also to develop tailored interventions that improve clinical outcomes for patient subgroups with severe illnesses. But participation in research is imperative to ensure benefits from research. Our study findings highlight the need to actively engage and recruit severely ill individuals. It is also important to consider factors that support retention as psychiatric genetic discoveries progress towards clinical applications-discovery studies generally require a single contact whereas clinical genetic studies often need to be longitudinal. But there is a critical gap in our knowledge as to the why there are differences in initial participation as well as retention rates in individuals with schizophrenia. Future studies should be designed to facilitate this type of inquiry and must engage individuals with lived experience not just as research participants but as research partners and investigators. Approaches such as community based participatory research has demonstrated value in psychiatric services research and should be considered in psychiatric and mental health genetics research.

Psychiatric genetics research in general (not just schizophrenia genetics research) has a significant underrepresentation of individuals of non-European ancestry (Akinhanmi et al., 2018; Martin, Kanai, et al., 2019; Trubetskoy et al., 2022). Despite concerted efforts (such as through policy initiatives by the NIH), this underrepresentation continues to widen (Akinhanmi et al., 2018; Fatumo et al., 2022; Peterson et al., 2019). Research findings from the general population show that there is a (historically justified) lack of trust among Black individuals when it comes to genetics research participation (McQuillan et al., 2003; Murphy & Thompson, 2009). But there are no studies that have examined genetics research participation specifically in severely psychiatrically ill groups of non-European ancestry. A study that examined racial differences in participation for a genetics of nicotine dependence study showed that willing to participate was not a significant barrier once the racial minority groups were reached by targeted recruitment and engagement (Hartz et al., 2011). It is unknown if such targeted recruitment and engagement would suffice to improve participation of individuals who have a severe mental illness or those with more severe symptoms in general. Studies of race and ethnic differences in utility of genetic findings and ethical concerns of individuals with severe mental illnesses are scant; most of them predominantly including White individuals (Costain et al., 2014; Hippman et al., 2013; Moldovan et al., 2019). Our study findings demonstrate the gap in schizophrenia genetic studies, but extant literature shows that this extends to other mental illnesses such as bipolar disorder (Akinhanmi et al., 2018). Race is not a valid genetic variable, but it is a valid and important social variable that influences important mental health outcomes and as such deserves due consideration in the design stage itself. This is important not just to ensure that research benefits are equitably distributed, but is a necessity to accurately quantify genetic risk (Martin, Daly, et al., 2019).

Our study findings show a statistically significant association of genetic study participant by the type of antipsychotic medication at baseline-those who were on second generation antipsychotic medications alone were more likely to participate. Second generation medications are the first line treatment for schizophrenia and patients who are on second generation medications tend to be more treatment responsive or are likely to have fewer years since initial diagnosis (Domino & Swartz, 2008). Our study design and data does not allow for further inquiry into this but suggest the need for further investigation. Our study findings are also limited in that, the design allows us to only examine initial participation and not retention, which is critically important for clinical genetics studies. Nevertheless, our study highlights the need to understand the intersectional influences of clinical and sociodemographic factors in an individual’s decision to participate in a genetic study. The genetic study was optional in CATIE; this allowed us to examine factors influencing participation for schizophrenia patients, but such studies involving diagnosed individuals are critically warranted in other psychiatric disorders including major depressive, bipolar disorders and eating disorders among others.

## Data Availability

Data can be accessed with permission from NIH NDCT

