## Supplement for "Influences of race and clinical variables on psychiatric genetic research participation: Results from a schizophrenia sample"

**Supplementary Table 1: N of complete data for relevant variables across time points**

| Variables | | Timepoints | | | |
|---|---|---|---|---|---|
| | | **Four** | **Three** | **Two** | **One** |
| Insight | | 765 | 941 | 1117 | 1328 |
| Illness severity (patient) | | 777 | 949 | 1117 | 1335 |
| Illness severity (clinician) | | 775 | 951 | 1127 | 1335 |
| Depression | | 785 | 952 | 1124 | 1338 |
| PANSS | | 786 | 953 | 1128 | 1339 |
| Neurocognition* | | - | 539 | 824 | 1165 |
| General physical health | | 757 | 934 | 1113 | 1325 |
| General mental health | | 757 | 934 | 1113 | 1325 |
| Quality of life | | 764 | 939 | 1112 | 1324 |
| Competencies* | Understanding | - | 628 | 964 | 1253 |
| | Appreciation | - | 628 | 964 | 1253 |
| | Reasoning | - | 628 | 964 | 1253 |
| | Choice | - | 628 | 963 | 1253 |

*****These variables only have 3 timepoints for consistent measurements for most participants.

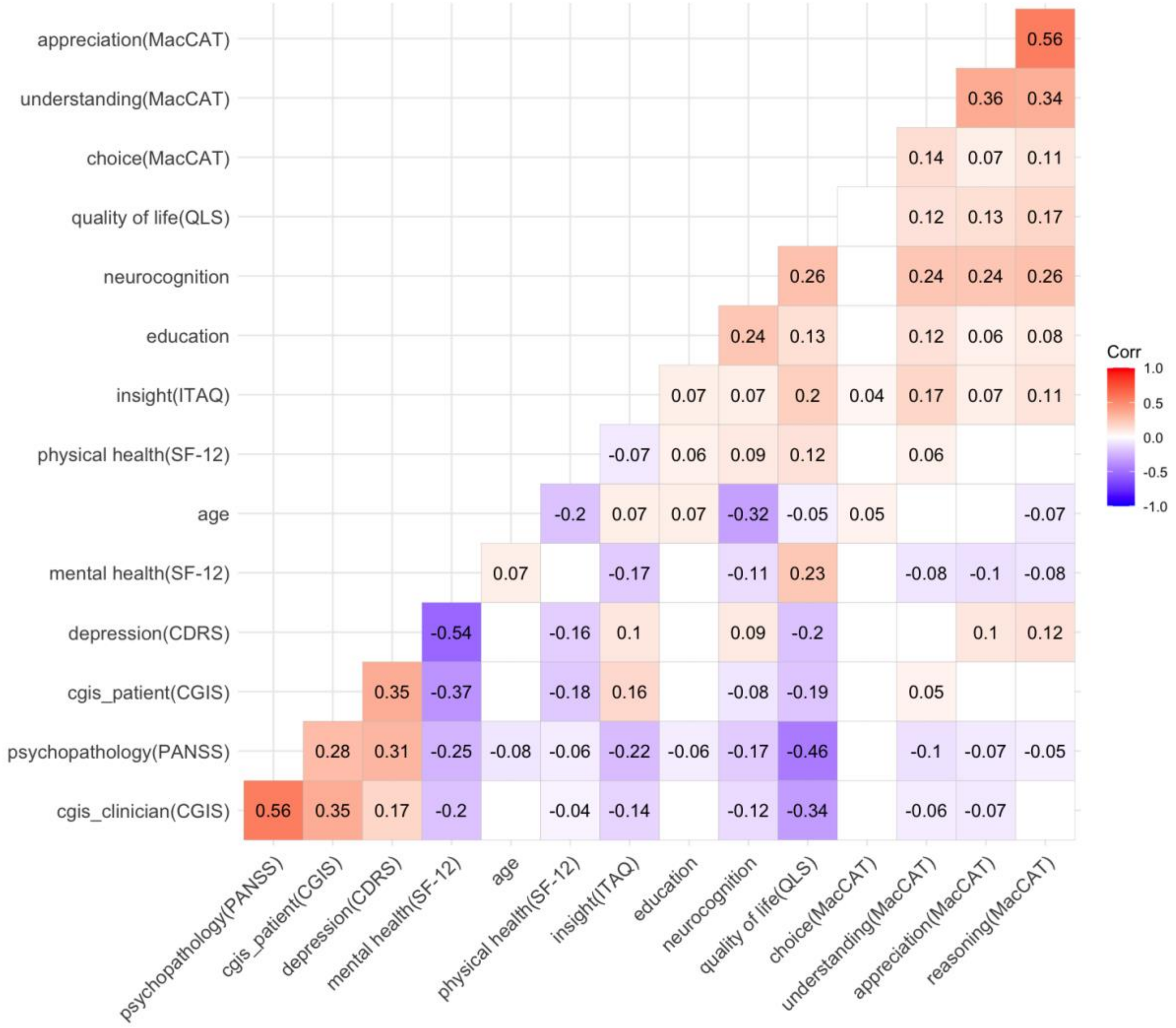


Supplementary Figure 1: Correlogram of relevant variables from baseline. Measures for variables are included in parenthesis. CDRS–Calgary Depression Rating Scale; CGIS–Clinical Global Impression of Symptom Severity; ITAQ–Illness and Treatment Attitude Questionnaire; MacCAT-CR–MacArthur Competence Assessment Tool- Clinical Research; PANSS–Positive and Negative Syndrome Scale; SF-12– 12-item Short Form Survey; QLS–Heinrich-Carpenter Quality of Life Scale.
